# Ukrainian syndrome. Youth in Ukraine in the face of a 2022-Full-scale Russian invasion

**DOI:** 10.1101/2023.01.08.23284304

**Authors:** Mykhailo Matiash, Spartak Subbota, Vitalii Lunov

## Abstract

Since 2014, Ukraine has been in constant military conflict. Every year hundreds of killed and wounded fighters, thousands of refugees, suffering of the civilian population in the battle zone. But a real large-scale disaster came to Ukraine on February 24, 2022, when a war began, which in terms of cruelty and the number of weapons used can only be compared with the Second World War.

In fact, Ukrainian syndrome is the result of any mental trauma: war, natural disaster, accident, mutilation, violence (sexual or domestic). The Ukrainian syndrome is a certain behavioral pattern of adaptation and an individual position of a Ukrainian regarding the Russian-Ukrainian war.

**Materials and methods:** The Scale of non-clinical assessment of behavioral changes and social functioning (block A, B. Rate on a scale: -1 means this sign used to be characteristic of me, but now it has disappeared, or it does not bother me; 0 means I practically do not feel it, or it is absent; 1 means there are certain signs, but they do not significantly affect life and well-being; 2 means the manifestation of this sign is strong; 3 means I feel exhausted and disorganized), and the Questionnaire of the experience of being in a combat zone developed by Vitalii Lunov were used in the study. The study was conducted from April 2022 to January 2023.

4292 youth and young scientists aged 16 to 40 from 61 cities of Ukraine took part in the research.

**Conclusions:** In this article, we have presented only the results of the descriptive statistics of the first part of the study of the Ukrainian syndrome, which reveal the prospects for building models of the rehabilitation potential of youth affected by the long-term Russian-Ukrainian war (Subbota, 2021).

The logic of the study provided for the objectification of the impact of direct military operations and the current situation on the life, material and property status and health of the subjects. In this context, we proceeded from the model of the spatial-relational disposition of a traumatic event (Lunov, 2022), which reflects the degree of objectivity of the loss / threat at the individual, family-related levels, and the level of the immediate environment.

Thus, it is worth noting the tendency of the field of property losses in the direction of their increase from the individual level to the level of the nearest social environment. This may be one of the factors inducing social stress as the ratio of personal losses to the losses of others. Significantly large losses in the immediate environment of an individual can act as potentially significant factors in reducing the zone of personal safety.

A similar trend is observed in relation to disability, bodily injuries, physical injuries suffered because of hostilities.

In general, we note that the attention of psychologists needs to be understood to the patterns of the reduction of the influence of symptoms present before the war and the appearance of new ones during the period of adaptation to the active phase of the Russian-Ukrainian war.

The very idea that some of the pre-war symptoms and behavior patterns that could have hindered the adaptation of the respondents in the conditions of war and internal displacement have disappeared opens prospects for further research into the resourcefulness of youth.

However, we cannot ignore the emergence of new symptoms and behavioral changes, especially those that are severe and that leave young people feeling exhausted and disorganized.

## Introduction

Since 2014, Ukraine has been in constant military conflict. Every year - hundreds of killed and wounded fighters, thousands of refugees, suffering of the civilian population in the battle zone. But a real large-scale disaster came to Ukraine on February 24, 2022, when a war began, which in terms of cruelty and the number of weapons used can only be compared with the Second World War. Every Ukrainian, without exception, felt this war. Putin’s troops occupied a fifth of our territory, bombing and rocket attacks on Ukrainian cities and villages became commonplace. And what the Russian occupiers did in the captured settlements will become material for the new Nuremberg trial. Seven months of war have passed - and we already have millions of refugees, hundreds of thousands of victims of the occupation, tens of thousands of wounded fighters, as well as widows, orphans, disabled people… Most of these people, like soldiers returning from the combat zone, have various mental disorders, because all they experienced acute stressful situations.

We remember the expression “Vietnam syndrome”, which united people whose psyche was affected by the events of the Vietnam War. By analogy - “Afghan syndrome”, “Chechen syndrome”… And now the term “Ukrainian syndrome” appeared in the psychological community. It seems that the same processes are taking place in Ukraine, but scientists are increasingly pointing out the differences between this phenomenon and similar ones in previous years. But before talking about the specifics of the “Ukrainian syndrome”, it is worth talking about this mental disorder in more detail.

In fact, “Ukrainian syndrome” is the result of any mental trauma: war, natural disaster, accident, mutilation, violence (sexual or domestic). The Ukrainian syndrome is a certain behavioral pattern of adaptation and an individual position of a Ukrainian regarding the Russian-Ukrainian war. In this context, we proceeded from the model of the spatial-relational disposition of a traumatic event, which reflects the degree of objectivity of the loss / threat at the individual, family-related levels, and the level of the immediate environment (Lunov, 2022).

**Our research is aimed at** determining socio-behavioral patterns and ways of developing negative and positive symptoms among Ukrainian youth in the conditions of the Russian-Ukrainian war and the internal forced displacement caused by it.

## Materials and methods

The “Scale of non-clinical assessment of behavioral changes and social functioning” (block A, B. Rate on a scale: -1 - this sign used to be characteristic of me, but now it has disappeared, or it does not bother me; 0 - I practically do not feel it, or it is absent; 1 - there are certain signs, but they do not significantly affect life and well-being; 2 – the manifestation of this sign is strong; 3 – I feel exhausted and disorganized), and the “Questionnaire of the experience of being in a combat zone” developed by Vitalii Lunov were used in the study. The study was conducted from April 2022 to January 2023.

The research protocol was approved by the Department of General and Medical Psychology of the Bogomolets National Medical University (protocol No. 15 dated 31.03.2022). The survey of respondents was conducted on the platform https://forms.gle/U99akzToALh2mXLc9. All respondents gave their consent to use the results of the research for scientific purposes.

4292 youth and young scientists aged 16 to 40 from 61 cities of Ukraine took part in the research.

The research was carried out in accordance with the scientific topics and projects of the laboratory of the psychology of learning of the G.S. Kostyuk Institute of Psychology of the National Academy of Educational Sciences of Ukraine “The potential of genetic psychology in the study of the interaction of subjects in the educational space” (No. 0121U107603, 2021-2023), the department of general and medical psychology of the Bogomolets National Medical University “Methodological, clinical, applied aspects of psychological medicine and psychological practice” (No. 0120U100656, 2020-2022). The research was carried out with the support of the grant of the LLC “National Institute of Evidence-Based Psychotherapy” “Metacognitive psychology and evidence-based psychotherapy” (No. 2022-MPEBP, 2022-2024).

## Results and Discussion

According to the ***criterion of age***, the respondents are divided as follows:

16-17 years old – 14.4%; 18-20 years old – 24.4%; 21-25 years old – 25.6%; 26-30 years old – 18.4%; 31-35 years old – 9.0%; 36-40 years old - 5.2%.

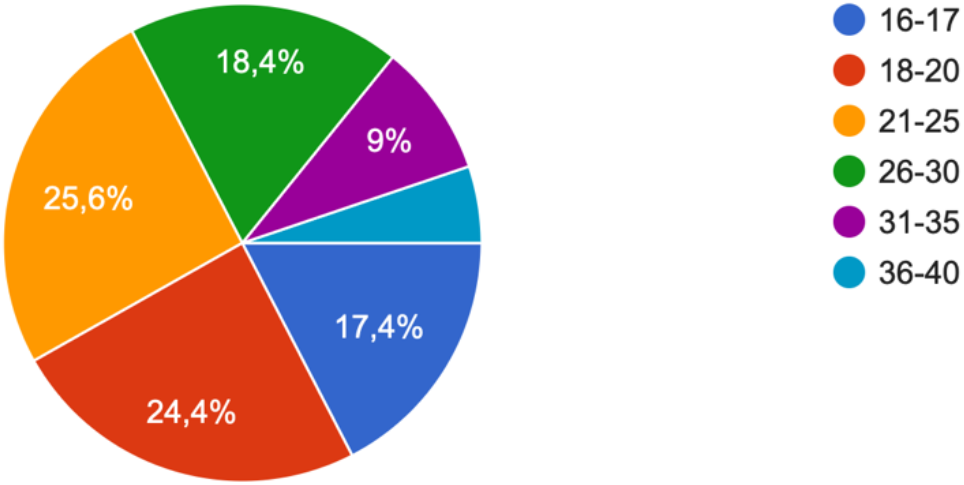

According to the ***criterion of gender***, the respondents are mostly representatives of the female gender - 89.3%, and 10.4% - male.

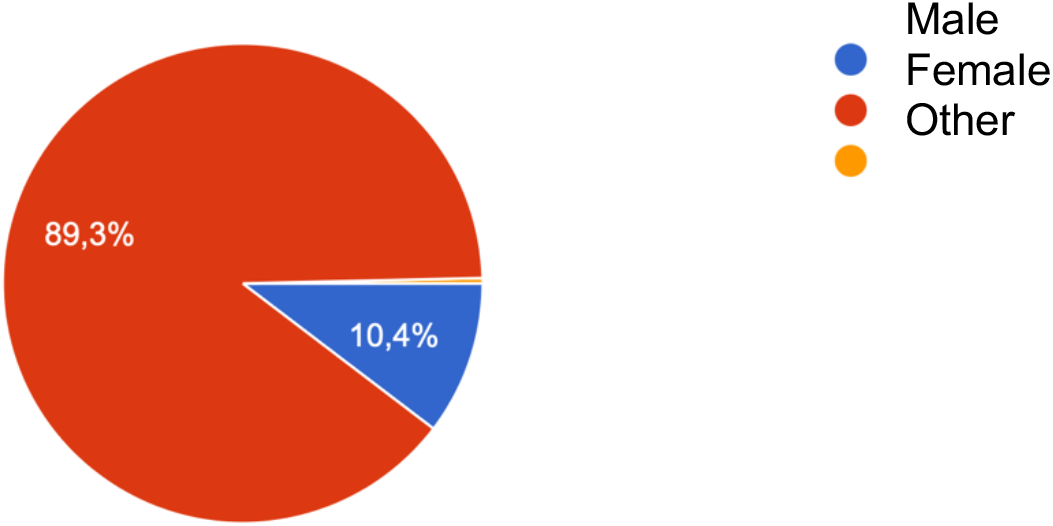

Regarding the question “***Have you been in a combat zone due to the Russian war in Ukraine (as of February 24, 2022)?***” respondents noted that 58.5% were not, 37.7% were in the war zone, and 3.8% are still.

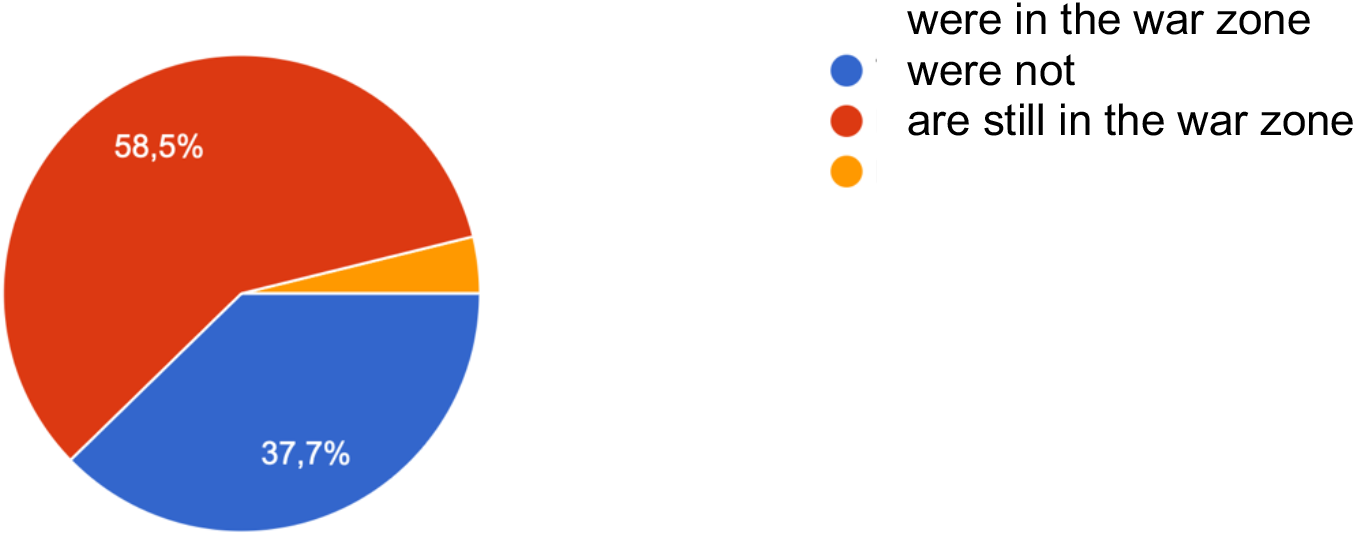

To the question “***Have you been in a combat zone before (before February 24, 2022)?***” respondents testified that 92.4% of them do not have such an experience, while 7.4% of respondents have such an experience in their lives.

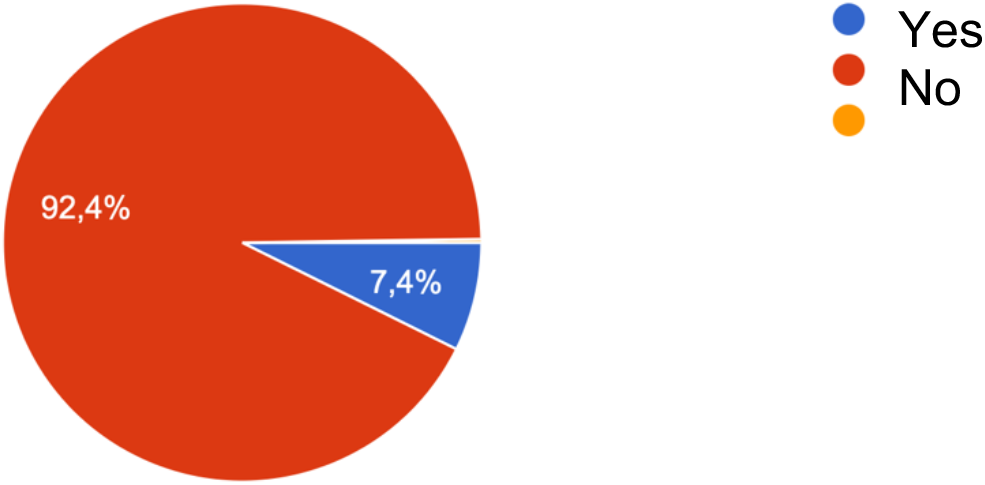

To the question “***Have you experienced destruction of housing, property, etc***.?” respondents indicated that 89.9% of them had not experienced this, while 10.1% had experienced home destruction.

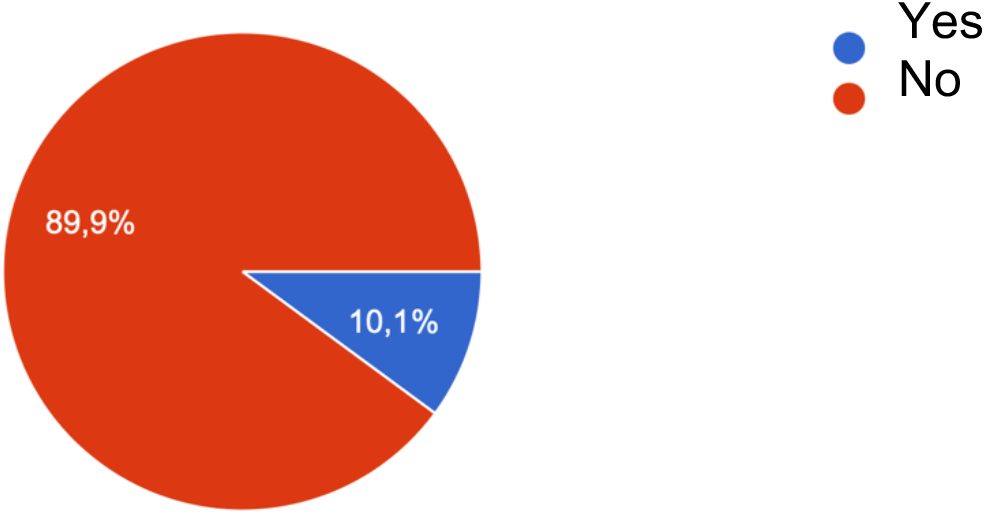

Regarding the question “***Have your relatives and family members experienced destruction of housing, property, etc.?***” the respondents indicated that 32.6% of them had relatives and family members who experienced property destruction, and 32.6% of the respondents did not have such experience with close relatives and family members.

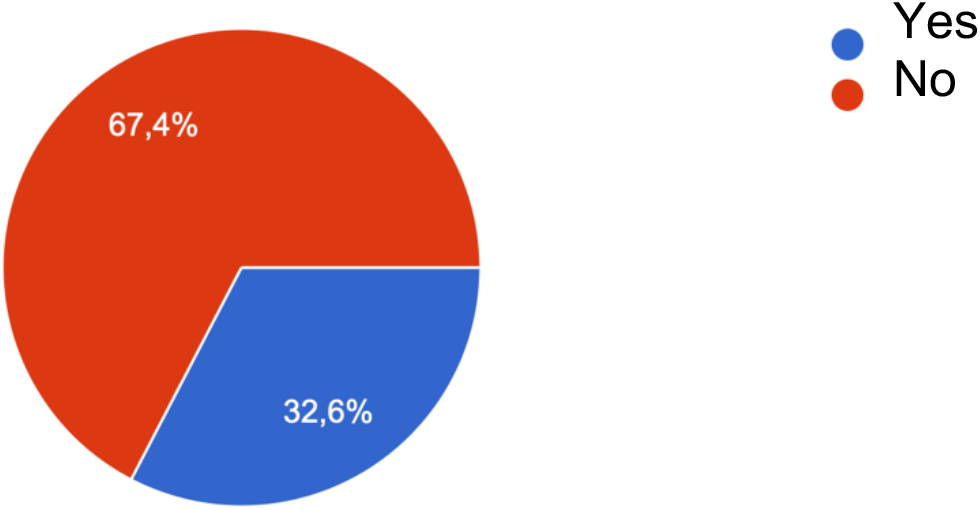

Answering the question “***Have people from your immediate environment experienced the destruction of housing, property, etc.?***” 39.4% of respondents talk about such experience, at the same time, 60.6% of respondents do not have such experience.

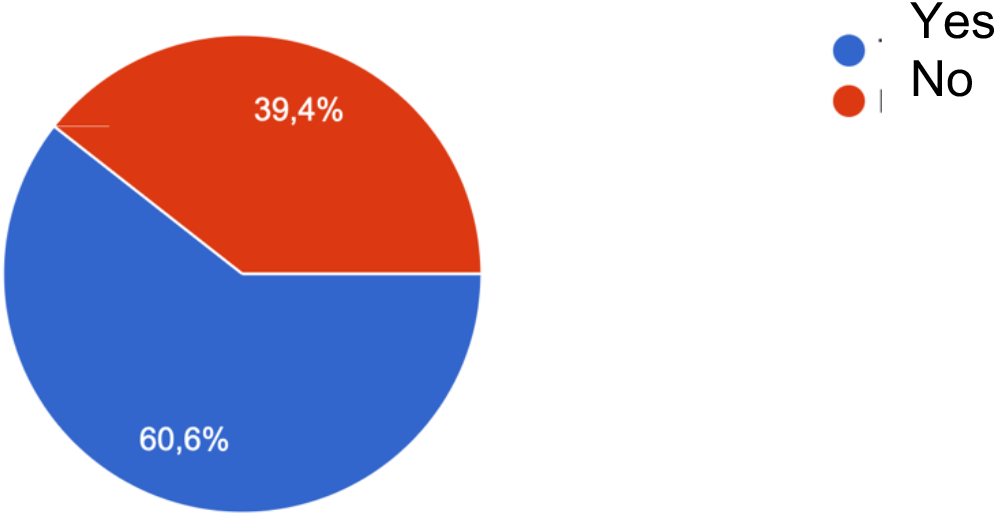

Regarding the question “***Have you suffered disability, bodily harm, physical injuries, etc. as a result of hostilities?***” only 0.7% of respondents claim such experience, while 99.3% do not have such experience.

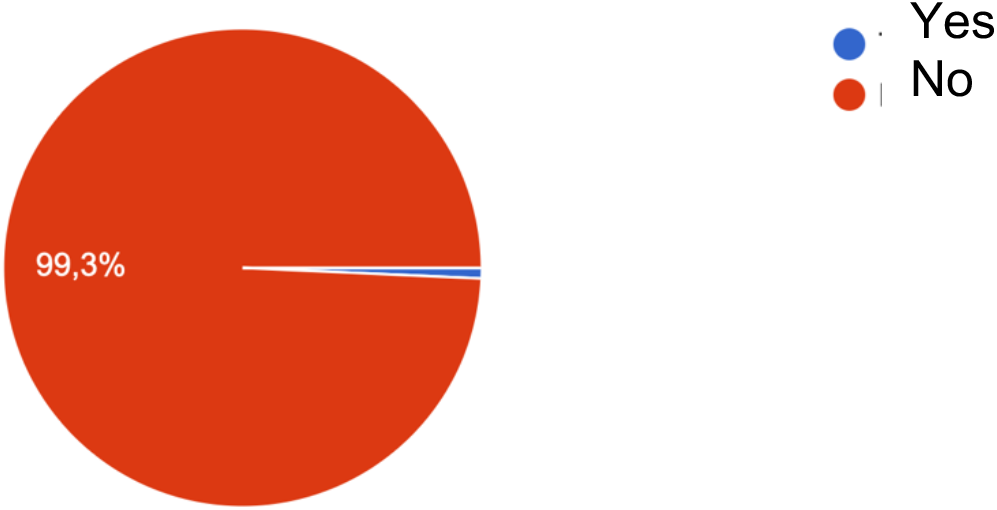

At the same time, to the question “***Have your relatives and family members been disabled, physically injured, physically injured, etc. as a result of hostilities?***” 15.7% of respondents indicate relevant experience, while 84.3% of respondents say they have no such experience.

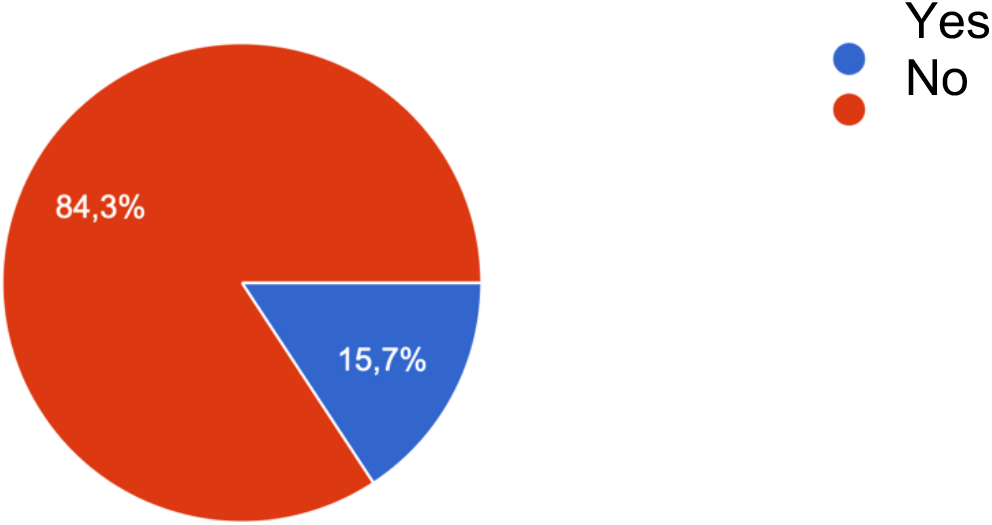

In addition, to the question “***Have people from your close environment experienced disability, bodily harm, physical injuries, etc. as a result of hostilities?***” 35.4% of the respondents answered about the presence of such a case, and 64.6% noted its absence.

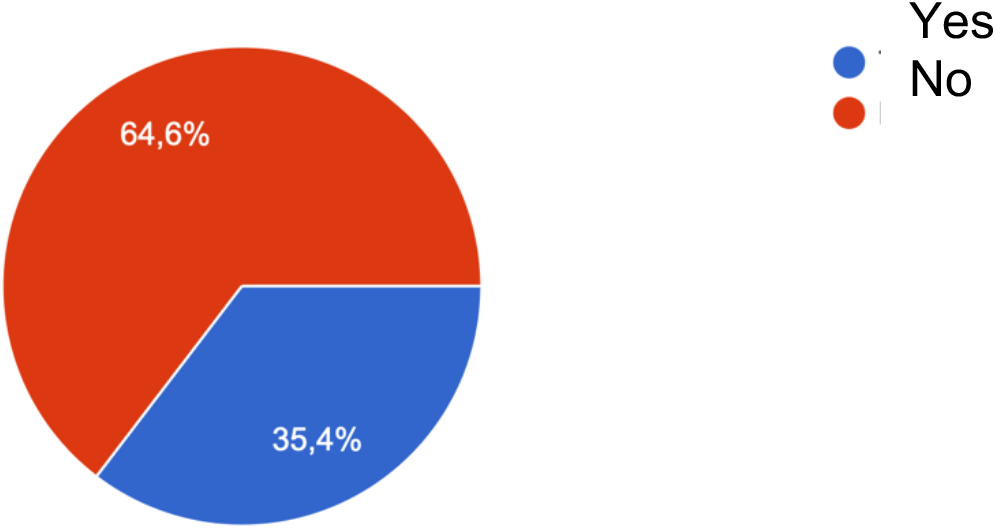

### The next block of questions is related to internal forced displacement and migration

To the question “***Have you been forced to leave your home in search of a safe place to live?***” 36.6% of respondents answered that they were forced to leave their homes; 41.8% - stayed at home; 19.2% initially left home but returned home; 13.2% do not have the opportunity to leave the war zone; and 2.3% constantly move from one city to another.

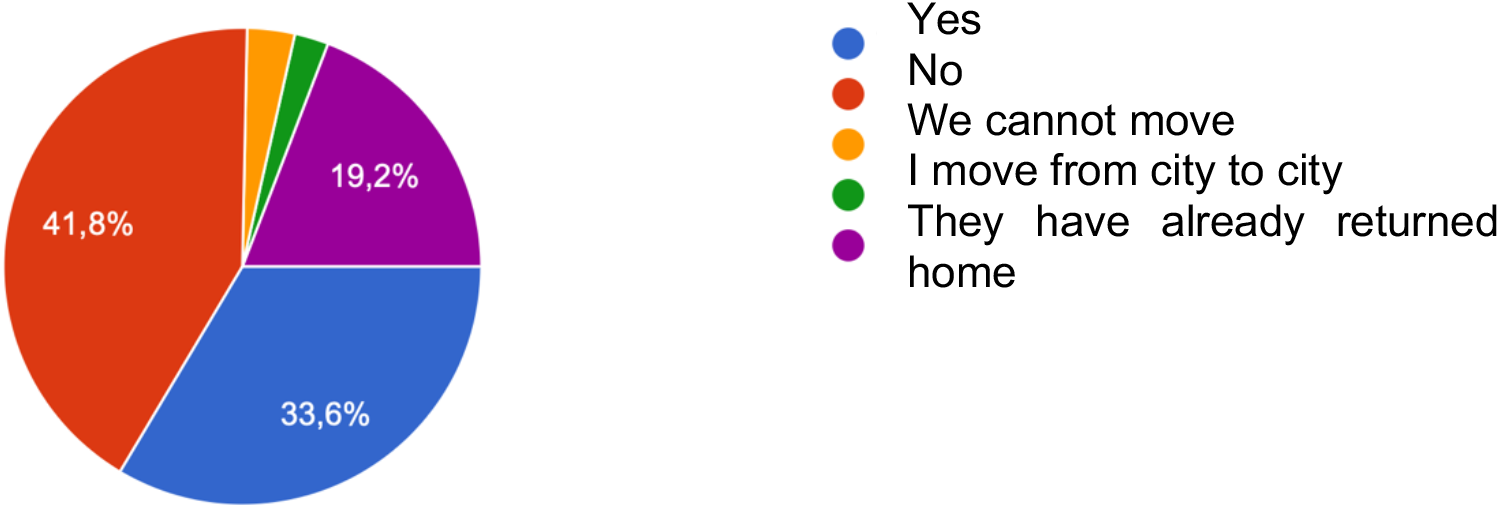

You should pay attention to the question “***Have your family members gone abroad?***”. In 33.3% of the studied young people, individual family members left the country. In 63.7%, all family members remained in Ukraine. Only 1.5% of respondents had their entire family move abroad. But 1.5% of respondents left this question unanswered.

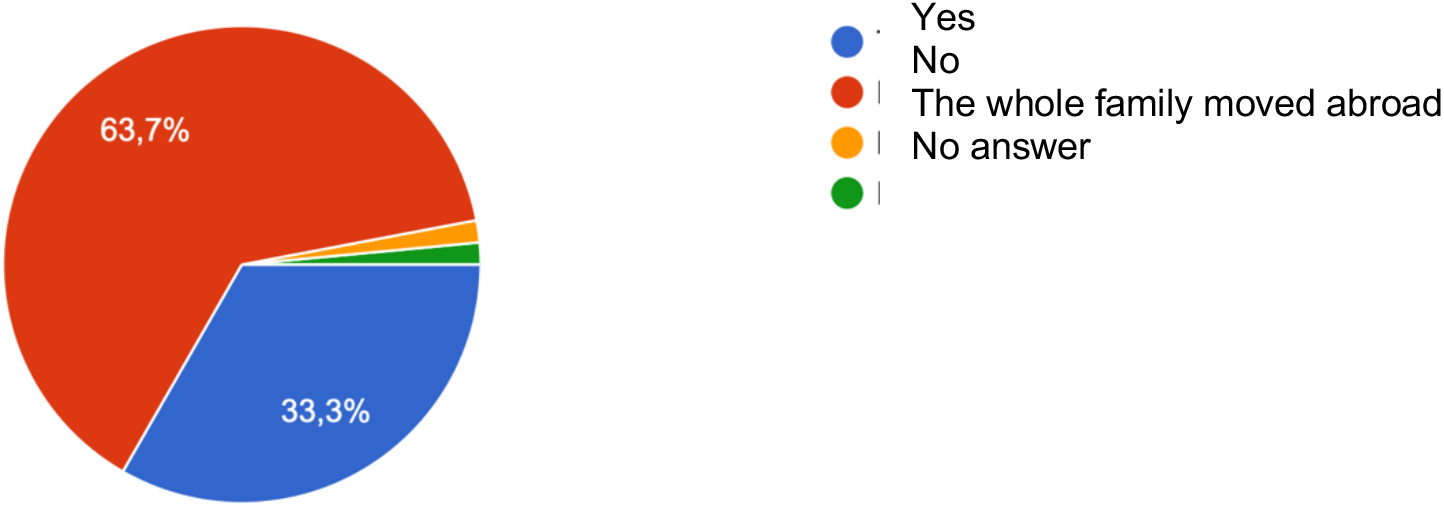

### The self-assessment of psychological, and behavioral changes by young people, which they experienced under the influence of the Russian-Ukrainian war during the period of full-scale Russian invasion from February 24, 2022

The self-assessment of psychological, and behavioral changes by young people, which they experienced under the influence of the Russian-Ukrainian war during the period of full-scale Russian invasion from February 24, 2022, deserves special attention in the study.

***To the question “I have new somatic (physical) symptoms”*** respondents noted the following.

4.9% of respondents note that they used to have new somatic symptoms from time to time, but now they have disappeared or do not bother them.

36.4% of respondents practically do not experience new somatic symptoms.

35.0% of respondents have certain signs of new somatic symptoms, but they do not significantly affect life and well-being.

12.6% of respondents testify that the manifestation of new somatic symptoms is strong.

And 11.1% of respondents feel exhausted and disorganized due to the appearance of new somatic symptoms.

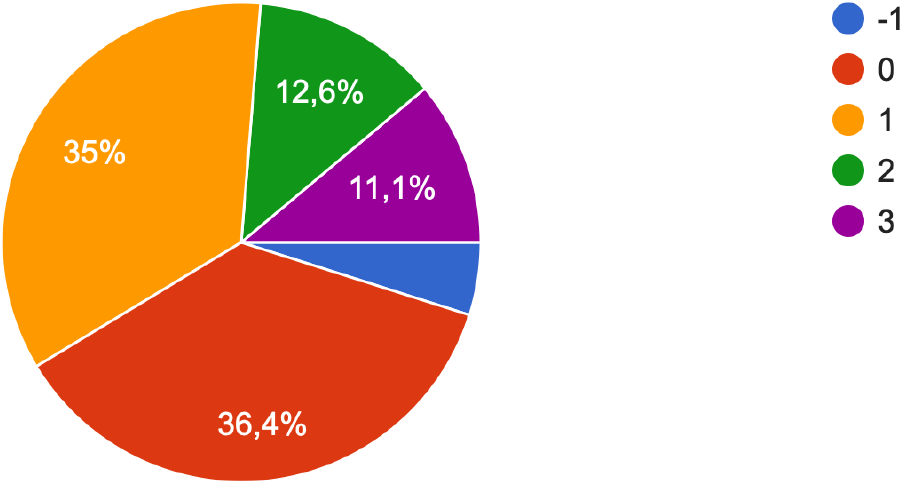

The next question was “***My late somatic (physical) symptoms have increased***”.

We note that 5.3% of the respondents noted that this feature used to be characteristic of them, but now it has disappeared or does not bother them. 47.6% of respondents practically do not experience an increase in late somatic symptoms. At the same time, 25.6% of respondents have certain signs of worsening of late somatic symptoms, but they do not significantly affect life and well-being. In 14.2%, the manifestation of late somatic symptoms is strong. And 7.3% of respondents feel exhausted and disorganized due to the influence of late somatic symptoms.

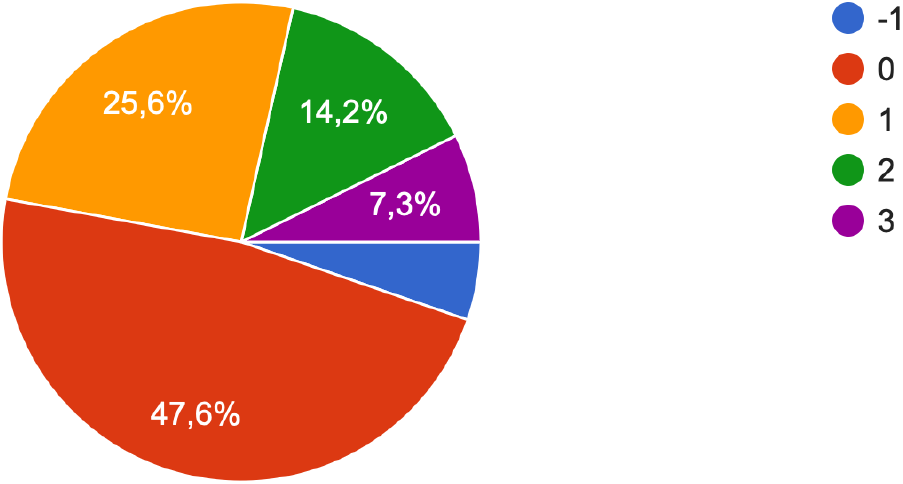

Regarding the question “***My late somatic (physical) symptoms have disappeared***” the following results were obtained.

9.6% of respondents note that their old somatic symptoms have disappeared.

69.7% of respondents also determine their complete absence.

13.3% of respondents say that their symptoms remain, but they do not significantly affect life and well-being.

4.2% of respondents say that the manifestation of late somatic (physical) symptoms is strong.

And 3.2% of respondents note that under the influence of these symptoms they feel exhausted and disorganized.

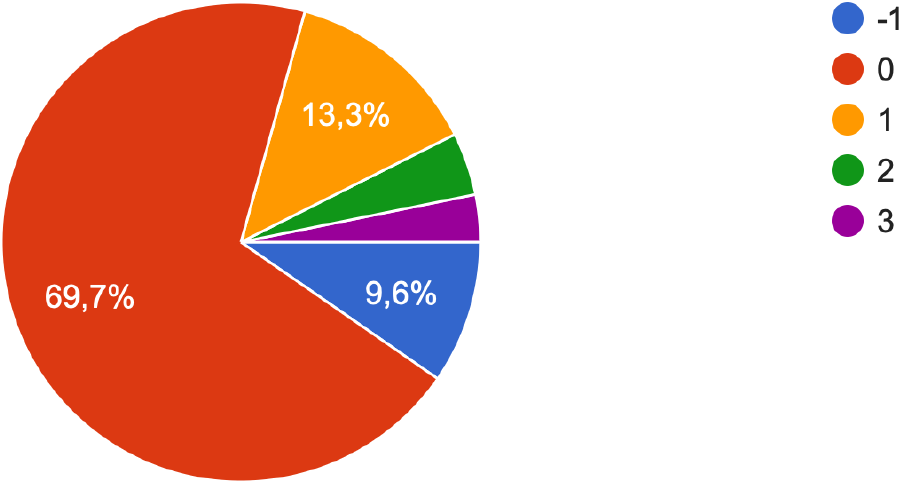

You should pay attention to the question “***Intrusiveness of thoughts, winding***”.

It was found that 7.3% of the respondents noted that this feature used to be characteristic, but now it has disappeared. 13.3% of respondents practically do not feel, or they do not have obsessive thoughts, winding. 36.9% of respondents in this question say that there are certain signs, but they do not significantly affect life and well-being. In 31.5%, the manifestation of this sign is strong. And 11.0 percent feel exhausted and disorganized due to obsessive thoughts.

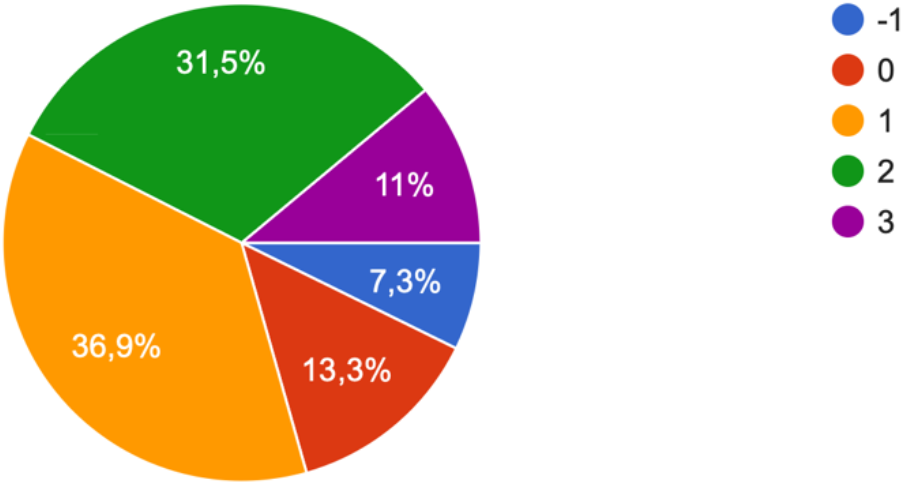

Let’s move on to the next question - “**Stupor**”.

Yes, 6.8% of respondents say that stupor used to be characteristic of them, but now it has disappeared or does not bother them.

44.9% of respondents practically do not feel a state of stupor.

31.9% have certain signs of stupor, but they do not significantly affect life and well-being.

12.9% of respondents testify that the manifestation of stupor is strong. But 3.5% of respondents say that they feel exhausted and disorganized due to stupor.

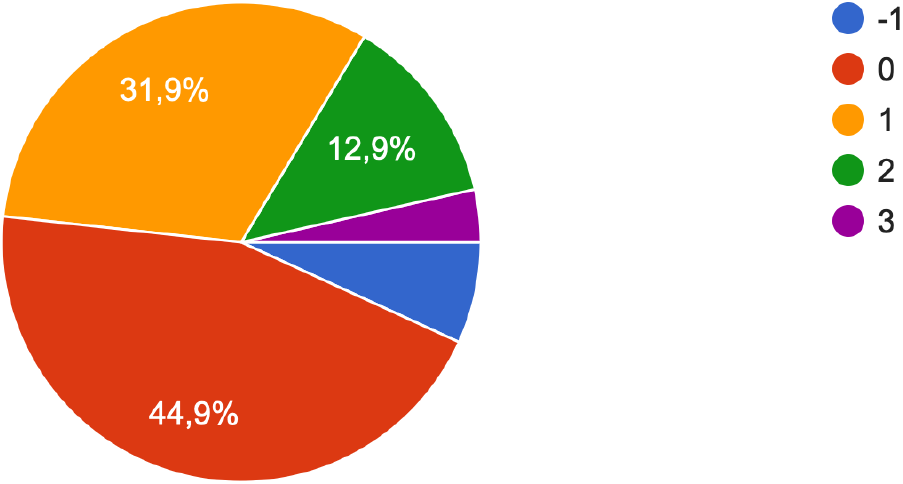

Let’s consider the answers to the question “***Compulsivity of actions and deeds***”. 5.7% of the respondents say that they used to be characterized by compulsive actions and deeds, but now they have disappeared or do not bother them. 51.5% of respondents practically do not feel the intrusiveness of actions and deeds. While 29.9% testify to certain symptoms, however, they do not significantly affect life and well-being. 10.3% of respondents say that the manifestation of this sign is strong. And 2.6% of respondents feel exhausted and disorganized due to the compulsiveness of actions and deeds.

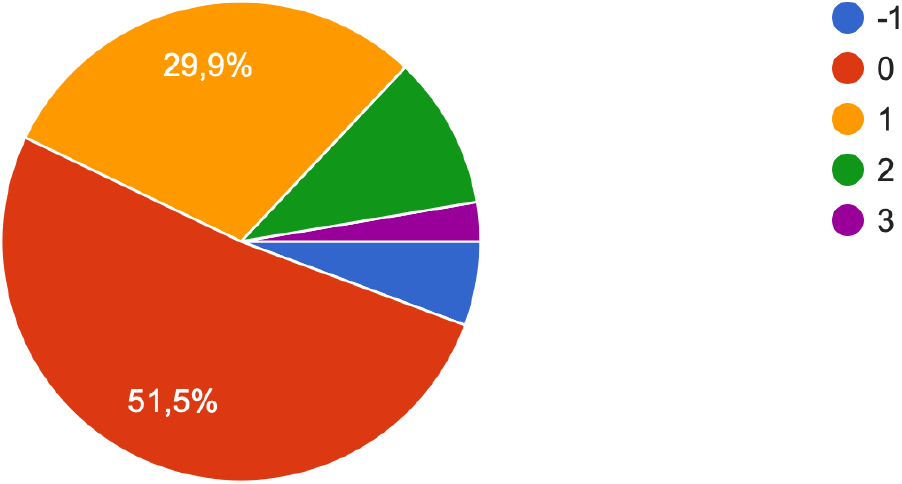

It is worth paying attention to the experience of ***hostility*** by Ukrainian youth.

Thus, according to the study, 2.3% of respondents got rid of the feeling of hostility that existed in the past. 24.4% of respondents practically do not feel it. 39.1% of respondents reported certain symptoms, but they do not significantly affect life and well-being. At the same time, 29.7% of respondents say that the manifestation of hostility in them is strong. And 4.5% of respondents testify that they feel exhausted and disorganized due to the feeling of hostility.

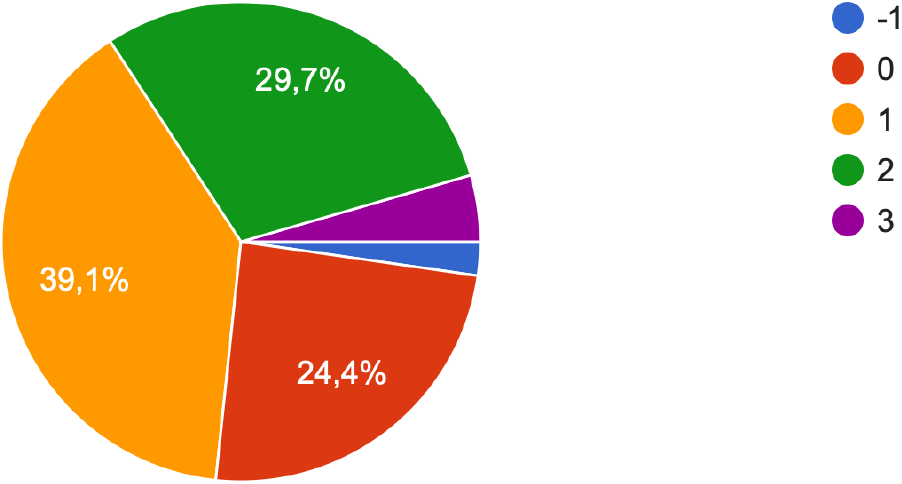

Regarding the self-assessment of the state of “***Phobia***” the following is established.

6.0% of respondents testify that phobias used to be characteristic of them, but now they have disappeared or do not bother them.

39.8% of respondents practically do not experience phobias. However, 35.2% have certain signs of phobias, but they do not significantly affect life and well-being. 15.4% of respondents have strong phobias. 3.7% of respondents feel exhausted and disorganized due to the manifestation of phobias.

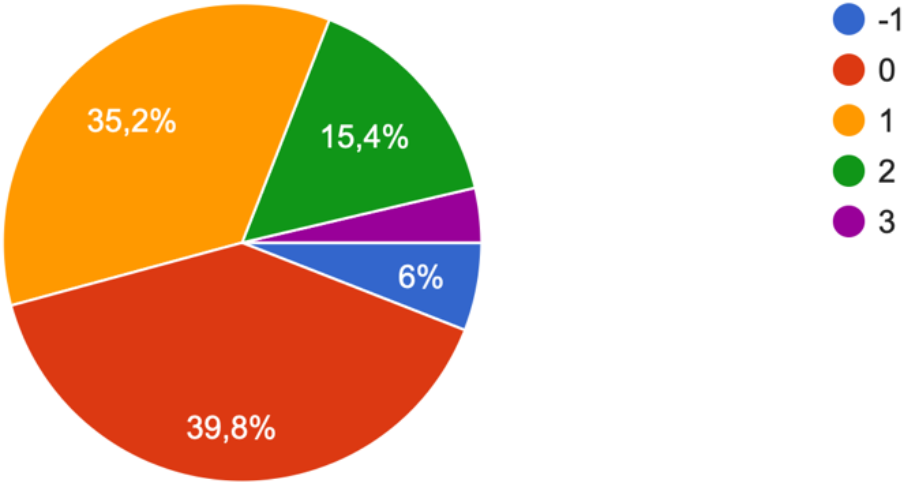

Regarding the self-assessment of the state of “***Anxiety***” the following results were obtained.

5.5% of respondents state that anxiety used to be characteristic of them, but now it has disappeared or does not bother them. 10.0% practically do not feel a state of anxiety. 35.2% of respondents observe certain signs of anxiety in themselves, but they do not significantly affect their life and well-being. At the same time, 33.8% of respondents show anxiety and testify that its manifestation is strong. At the same time, 15.4% said that anxiety makes them feel exhausted and disorganized.

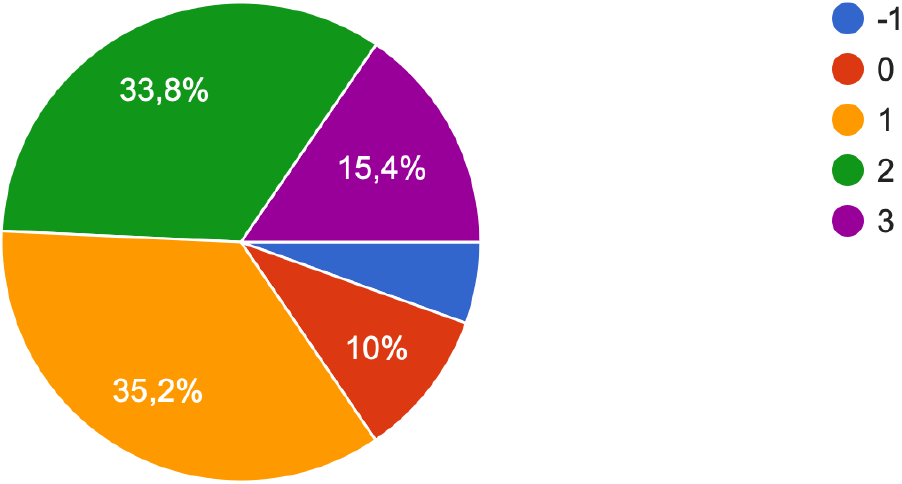

It is worth paying attention to the results of the self-assessment of the question “***Depressive state***” in respondents.

6.6% of respondents say that the depressive state used to be characteristic of them, but now it has disappeared or does not bother them. Also, 26.6% practically do not feel it, or it is absent. 34.3% of respondents testify that they feel certain signs of depression, but they do not significantly affect their life and well-being. At the same time, 21.7% of respondents say that the manifestation of a depressive state is strong in them, and 10.8% feel exhausted and disorganized due to a depressive state.

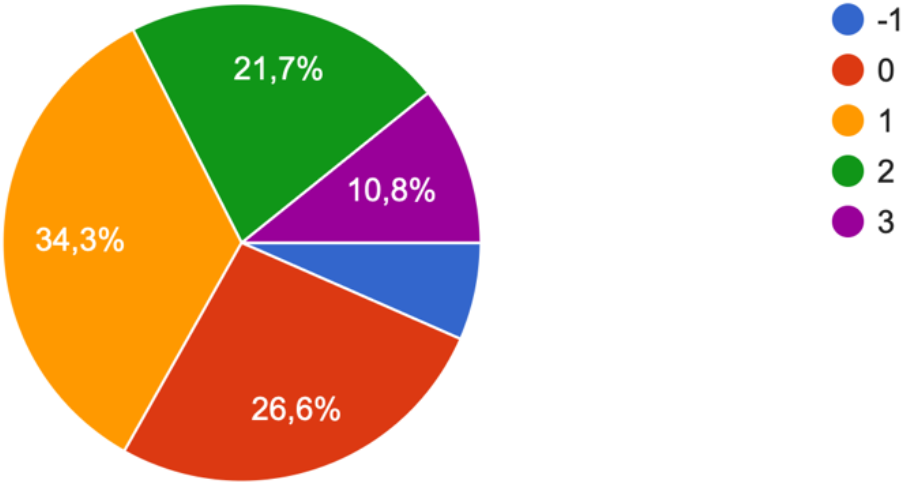

Regarding the assessment of the problem of “***Insomnia***” respondents answered in the following way.

Thus, 6.5% of respondents noted that the problem of insomnia used to be characteristic of them, but now it has disappeared or does not bother them. 39.0% of respondents practically do not experience the problem of insomnia, it is absent. 27.7% have certain symptoms of insomnia, but they do not significantly affect life and well-being. At the same time, 19.5% say that the manifestation of insomnia is strong, and 7.4% of young people say that they feel exhausted and disorganized because of insomnia.

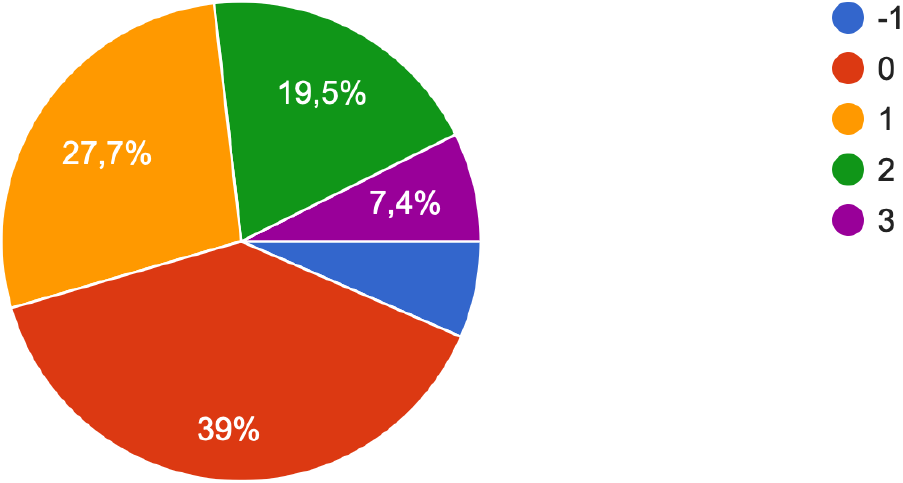

Regarding “***Problems falling asleep***” the following results were obtained.

5.7% of respondents note that earlier the problem with falling asleep was peculiar to them, but now it has disappeared or does not bother them. 31.3% of respondents practically do not feel such a problem. At the same time, 27.9% say that there are certain signs of problems with falling asleep, but they do not significantly affect life and well-being. In 24.6% of respondents, the manifestation of the problem with falling asleep is strong. And 10.6% of respondents feel exhausted and disorganized due to problems with falling asleep.

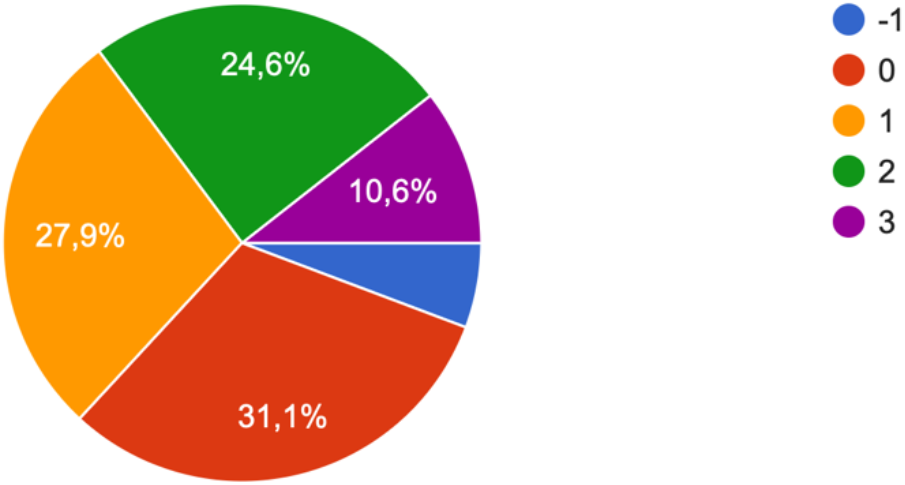

Analyzing the answers to the question “***Problems with the stability of sleep***” the following can be defined.

It was found that 3.4% of the respondents previously had problems with the stability of sleep, but now this problem has disappeared or does not bother them. 29.1% of respondents practically do not feel this problem. Also, 28.8% of respondents say that there are certain signs of sleep stability problems, but they do not significantly affect life and well-being. At the same time, 27.3% of respondents feel that the manifestation of the problem of sleep stability is strong. And 11.3% demonstrate that they feel exhausted and disorganized due to problems with the stability of sleep.

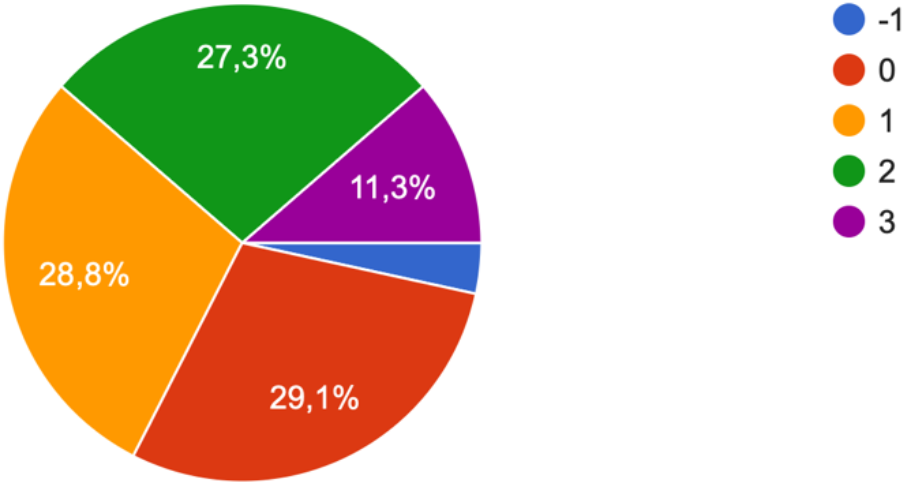

Regarding the respondents’ self-assessment of the question “***Nightmares***” the following results were obtained.

7.1% of respondents state that they used to have nightmares during sleep, but now they have disappeared or do not bother them. For 52.4% of respondents, this problem is practically not felt, or is absent. 25.5% of respondents say that there are certain symptoms, but they do not significantly affect life and well-being. Along with the fact that it deserves attention, 11.6% of respondents say that the manifestation of this feature is strong. And 3.3% of respondents feel exhausted and disorganized due to nightmares.

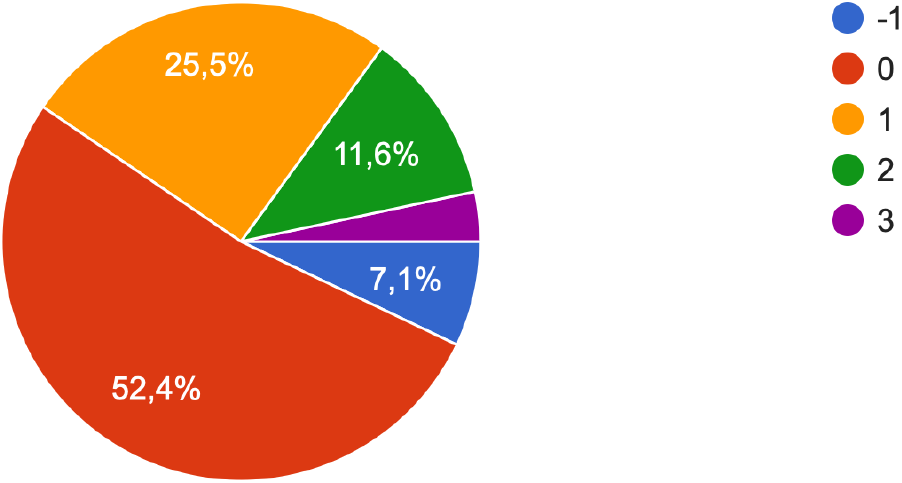

Considering “***Problems with awakening from sleep***” the following was established among the respondents.

3.5% of respondents say that I used to have problems waking up, but now they have disappeared or do not bother me. 33.4% have practically no problems waking up from sleep. 29.6% demonstrate that there are certain signs, but they do not significantly affect life and well-being. While 23.3% of respondents report that this symptom is severe, 10.2% of respondents reported feeling tired and disorganized due to problems waking up from sleep.

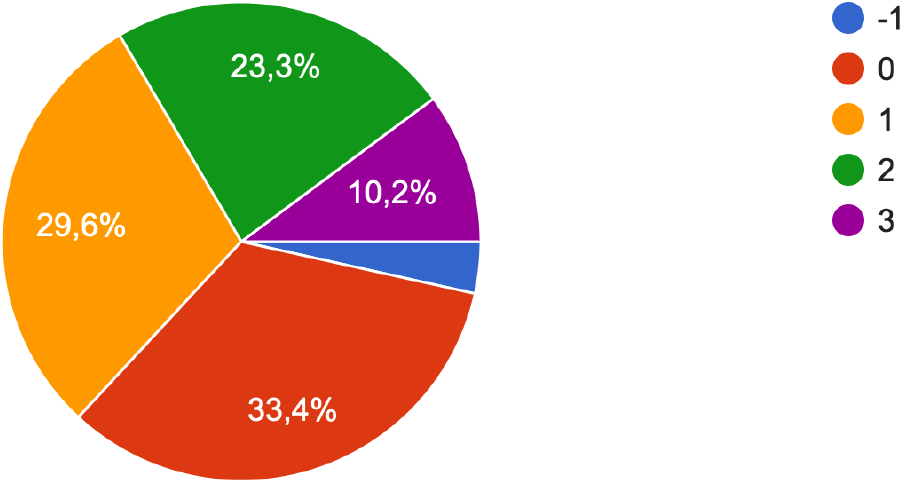

### Ukrainian youth’ social skills and functioning in relations

The next block of questions was related to respondents’ self-analysis of their social skills during the war. To the question “***I have lost my social skills and functions***” the following responses were received from the respondents.

6.3% of respondents indicate that they lost social skills and functions in the past, but not now. 51.6% of respondents testify that they practically do not feel the loss of social functions and skills. However, 24.6% have certain signs of loss of these functions, but they do not significantly affect life and well-being. It is worth noting that 13.4% of respondents note significant loss of social functions, and 4.0% feel exhausted and disorganized because of this.

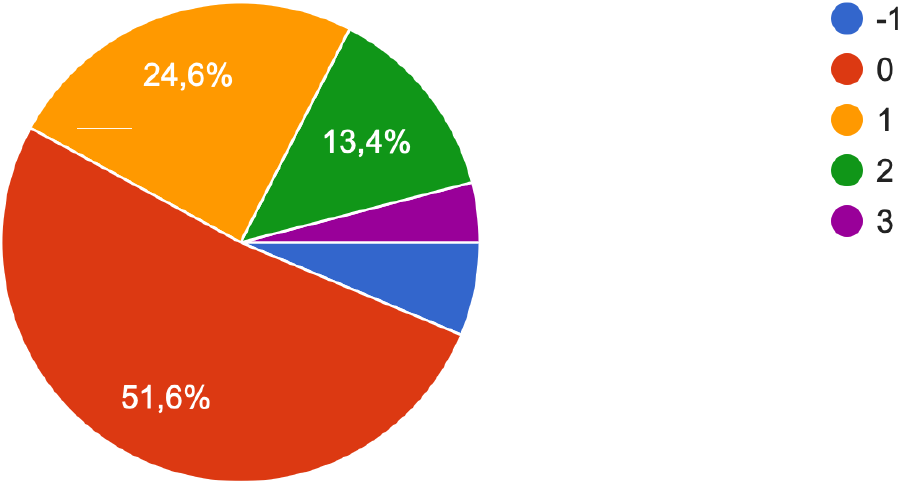

Regarding the acquisition of new social skills and functions by Ukrainian youth affected by the Russian-Ukrainian war, the following was found.

Answering the question “***I have acquired new social skills and functions***” the respondents gave the following answers.

4.5% of respondents find that they used to be able to acquire new skills, but now there are problems with it. 39.1% of respondents practically do not feel the emergence of new skills in them. At the same time, 33.9% of respondents demonstrate that they are developing new social skills, but they do not significantly affect their life and well-being. In addition, 21.0% of respondents note the confident mastery of new social functions and skills in their lives. While the need to develop these functions in 1.4% of respondents leads to a feeling of exhaustion and disorganization.

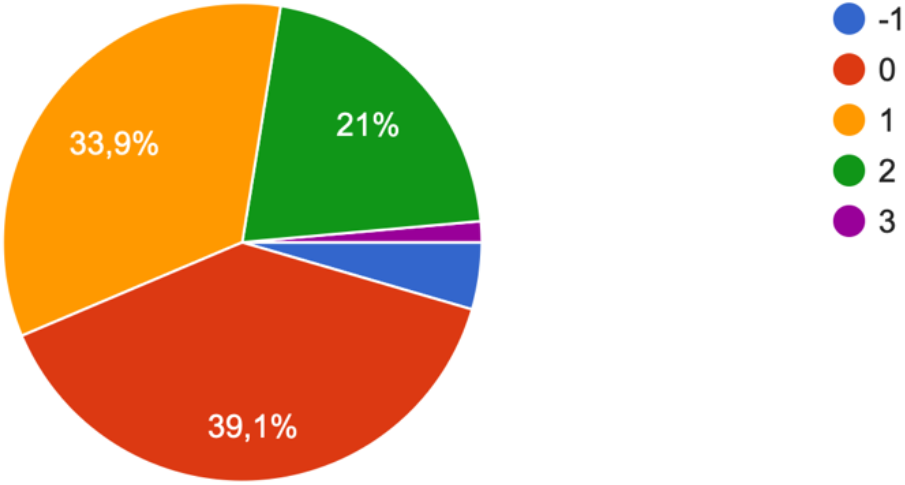

Regarding the question “***Conflicts with family members***” the respondents answered as follows.

8.2% of respondents noted the improvement of family relations. 31.0% of respondents find that they practically do not feel the presence of conflicts with family members, or they are absent. At the same time, 33.4% of respondents testify that there are conflicts in the family, but they do not significantly affect life and well-being. However, 21.0% of respondents have a strong manifestation of conflicts in the family, and 6.5% feel exhausted and disorganized due to conflicts.

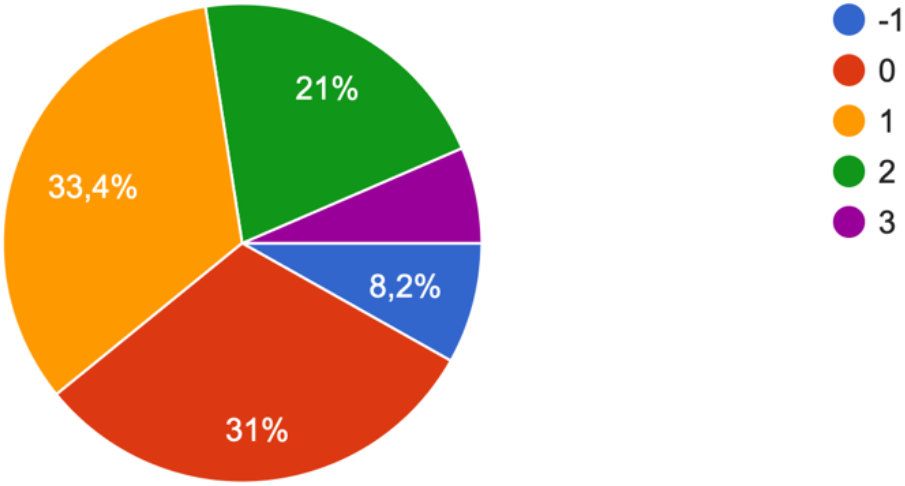

Regarding the respondents’ self-assessment of the question “***Conflicts with friends***” the following is summarized.

6.0 % of respondents say that they got rid of past conflicts with friends. 61.5% currently have practically no conflicts with friends and do not feel them. At the same time, 23.3% testify that there are certain signs of conflict with friends, but they do not significantly affect life and well-being. However, 7.6% of respondents have a strong manifestation of conflicts, and 1.6% feel exhausted and disorganized due to conflicts with friends.

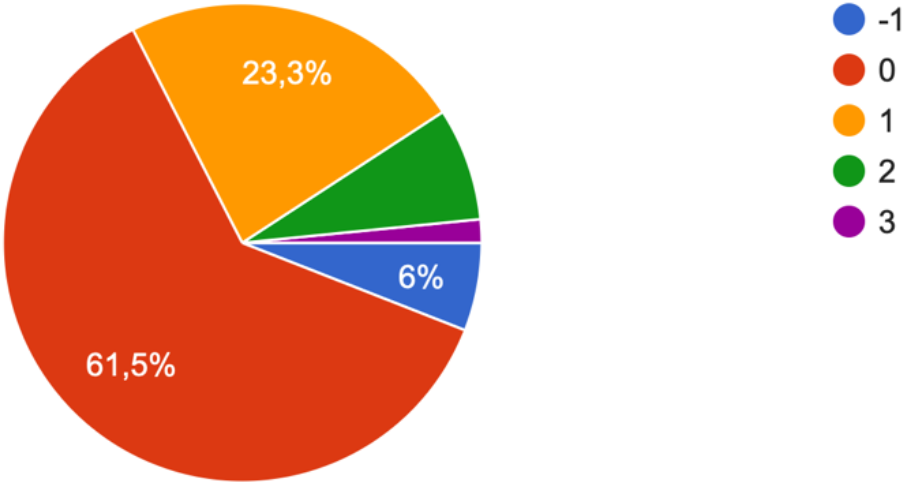

And finally, to the question “***Conflicts with colleagues***” the following was found among Ukrainian youth.

6.1 % of respondents claim that conflicts with colleagues used to be typical for them, but now they have disappeared or do not bother them. 72.8% of respondents practically do not feel conflict, or it is absent at all. 15.5% of respondents have certain signs of conflict with colleagues, but they do not significantly affect life and well-being. However, only 4.6% say that there is a strong conflict, and 1.0% feel exhausted and disorganized due to a conflict with colleagues.

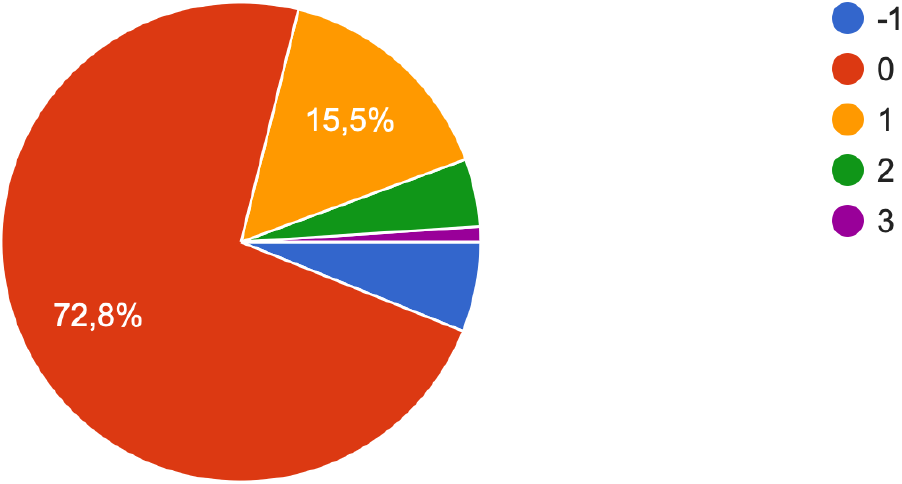

## Conclusions

In this article, we have presented only the results of the descriptive statistics of the first part of the study of the Ukrainian syndrome, which reveal the prospects for building models of the rehabilitation potential of youth affected by the long-term Russian-Ukrainian war (Subbota, 2021).

## Data Availability

All data produced in the present study are available upon reasonable request to the authors

https://forms.gle/U99akzToALh2mXLc9

## Declaration of Conflicting Interests

The author(s) declared no potential conflicts of interest with respect to the research, authorship, and/or publication of this article.

**Ethics committee of G.S. Kostyuk Institute of Psychology of the National Academy of Educational Sciences and Department of General and Medical Psychology of the Bogomolets National Medical University of Ukraine (protocol No. 15 dated 31.03.2022) gave ethical approvals for this work**.

